# Nonfasting, Telehealth-Ready LDL-C Testing With Machine Learning to Improve Cardiovascular Access and Equity

**DOI:** 10.1101/2025.10.27.25338909

**Authors:** Ronald Doku, Nana Yaw Osafo, John Kwagyan, William M. Southerland

**Affiliations:** Howard University College of Medicine, Washington, DC, USA

**Keywords:** low-density lipoprotein cholesterol, cardiovascular quality improvement, healthcare delivery, machine learning, non-fasting lipid panel, telehealth, health equity, value-based care

## Abstract

**Importance:** Current LDL-C testing requires 9–12 hour fasting and in-person visits, creating an access crisis: 40% of lipid panels occur outside fasting windows (yielding unreliable results), 60% of US counties lack cardiology services, and millions of patients with diabetes cannot safely fast. Meanwhile, telehealth infrastructure expanded 38-fold post-COVID, yet lipid workflows remain anchored to 1970s protocols. This mismatch drives ~ 20 million unnecessary repeat visits annually, disproportionately burdening Medicaid populations, essential workers, and rural communities.

**Objective:** To demonstrate that machine learning can transform lipid testing from a fasting-dependent, clinic-based bottleneck into an accurate, equitable, telehealth-ready service—eliminating three structural barriers simultaneously: fasting requirements, in-person visits, and racial algorithmic bias.

**Design, Setting, and Participants:** Cross-sectional analysis of All of Us Research Program (n=3,477; test n=696). Crucially, 40.1% were tested outside traditional fasting windows, reflecting real-world practice. We evaluated performance stratified by fasting status, telehealth feasibility (labs-only configuration), racial equity metrics, and economic impact.

**Main Outcomes and Measures:** Primary: MAE and calibration in non-fasting states. Secondary: Labs-only non-inferiority (±0.5 mg dL^−1^margin); racial equity (Black-White performance gap); economic savings from eliminated repeat visits; and classification accuracy at treatment thresholds (70, 100, 130 mg dL^−1^).

**Results:** The ML system demonstrated paradoxical superiority in non-fasting conditions—precisely when needed most. While equations deteriorated (Friedewald MAE 29.1 vs 25.9 mg dL^−1^fasting, slopes 0.58–0.61), ML maintained accuracy (24.0 vs 23.2 mg dL^−1^, slopes 0.99–1.07), achieving 17.2% improvement over Friedewald when non-fasting vs 10.4% fasting. Labs-only configuration proved non-inferior (MAE=-0.12, p<0.001), enabling national retail-pharmacy and home-testing workflows. The system achieved racial equity without race input (Black-White gap −0.19 mg dL^−1^, CI includes zero) while providing greatest improvement for Black patients (19% vs 11% for White). Economically, eliminating 4,000 repeat visits per 10,000 tests helps address an estimated $2 billion annual repeat-testing cost burden and yields $815,000 total savings per 10,000 tests ($245,000 direct healthcare, $570,000 patient costs), with break-even at just 750 tests.

**Conclusions and Relevance:** This ML approach helps address an estimated $2 billion annual problem of repeat testing while tackling three critical quality gaps in cardiovascular prevention: delayed treatment initiation, poor monitoring adherence, and specialty access barriers. By enabling accurate non-fasting, telehealth-compatible, race-free LDL-C estimation, it transforms lipid testing from an access barrier into an access enabler—particularly for the Medicaid, Medicare Advantage, and rural populations who drive both cost and outcomes in value-based care. From a technical standpoint, implementation requires only routine labs and <100 ms computation, making deployment feasible with existing infrastructure.

## 1 Introduction

### 1.1 The Access Crisis in Lipid Testing

Cardiovascular disease remains the leading cause of death in the United States, with lipid management central to prevention.[1] Yet access to accurate LDL-C testing is profoundly inequitable. Traditional testing requires 9–12 hour overnight fasting and in-person blood draws, turning a simple lab test into a series of cascading barriers:

#### Geographic barriers

60% of US counties lack cardiology services; rural patients travel a median of 40 miles for specialty lipid clinics.[2] Fasting requirements necessitate early morning appointments, often requiring overnight stays or 3–4 hour pre-dawn drives.

#### Socioeconomic barriers

Shift workers cannot easily accommodate fasting windows or morning appointments. Caregivers (predominantly women) struggle with childcare during testing. Hourly workers lose wages for clinic visits.[3]

#### Medical barriers

Patients with diabetes face hypoglycemia risk during prolonged fasting. Those on insulin cannot safely skip meals. Multiple comorbidities require more frequent monitoring, multiplying access challenges.[4]

#### The scale of the problem

In our cohort, 40.1% were tested outside traditional fasting windows—representing either protocol violations or practical accommodations. Nationally, this suggests roughly 20 million annual non-fasting lipid panels in suboptimal conditions, precisely where equation-based LDL-C estimates are most likely to fail.

### 1.2 The Telehealth Opportunity

The COVID-19 pandemic catalyzed explosive telehealth growth: virtual visits increased 154-fold (0.1% to 15.4% of encounters).[5] Home phlebotomy services expanded. Retail pharmacies added point-of-care testing. Yet lipid management hasn’t kept pace because:

- Standard equations assume fasting and often lose accuracy in non-fasting states [4].
- Blood pressure is not routinely available in retail or home testing settings.
- Race-based corrections in clinical algorithms raise equity concerns [6].

The 2016 European Atherosclerosis Society endorsed non-fasting lipids,[4] but US adoption lags due to accuracy concerns with calculated LDL-C in non-fasting states.

### 1.3 The Technical Challenge

Standard LDL-C equations (Friedewald, Martin-Hopkins, NIH Equation 2) were validated in fasting populations and degrade in non-fasting states—particularly at elevated triglycerides where postprandial lipemia compounds errors.[7] This creates a cruel paradox: the populations needing convenient testing most (diabetics, rural patients, shift workers) are precisely those where equation failures are most severe.

Recent machine learning approaches show promise[8] but haven’t demonstrated: (1) maintained accuracy across fasting states, (2) feasibility with labs-only data (no vitals), or (3) racial equity without race-based adjustments.

### 1.4 Study Objectives

We hypothesized that machine learning could maintain LDL-C accuracy in three access-enabling scenarios:

- **Non-fasting states:** Patients can be tested at any time, eliminating scheduling barriers.
- **Labs-only configuration:** No blood pressure required, enabling telehealth and retail models.
- **Race-free inputs with equity audit:** Equitable performance without race-based corrections.

We evaluated real-world performance, economic impact, and implementation pathways to inform access-expanding deployment.

### 1.5 Why This Moment Demands Change

The case for rethinking LDL-C workflows has reached a critical inflection point. Four converging forces make non-fasting, telehealth-ready testing not just desirable but essential:

#### 1. Telehealth infrastructure without matching workflows

Post-COVID, virtual visits increased 38-fold and stabilized as a permanent care modality.[5] Home phlebotomy services expanded nationwide. Retail pharmacies added point-of-care testing. Yet lipid protocols still require 9–12 hour fasting and in-person visits—a mismatch that blocks millions from accessing established telehealth infrastructure.

#### 2. Modern therapies creating new monitoring demands

Use of advanced diabetes therapies increased sharply from 2020-2023, and tens of millions of Americans now require frequent lipid monitoring but often cannot safely fast due to hypoglycemia risk. The very populations needing most monitoring face the highest barriers.

#### 3. Health system capacity crisis

With 60% of US counties lacking cardiology services and primary care facing critical workforce shortages, every unnecessary repeat visit represents lost capacity.[2] Rural clinics operating at maximum capacity cannot afford 40% of lipid panels requiring second visits for fasting compliance.

#### 4. Regulatory push for algorithmic equity

CMS and major payers now require equity metrics in quality programs. Race-based clinical algorithms are being systematically removed.[6] Health systems need tools that improve accuracy while demonstrating measurable equity gains—exactly what our race-free, ML-based approach provides.

Against this backdrop, maintaining fasting-dependent, in-person-only lipid testing is not neutral—it actively perpetuates access inequities and system inefficiencies at precisely the moment when flexible, equitable solutions are both technologically feasible and urgently needed.

## 2 Related Work

### 2.1 Non-fasting Lipid Testing and Guideline Evolution

For decades, routine lipid testing required 9–12 hours of fasting, in part because the Friedewald equation assumes fasting triglycerides (TG).[9] Accumulating evidence shows fasting is unnecessary for most patients. In a 2016 European consensus statement, Nordestgaard *et al*. reviewed large cohorts and recommended *non-fasting* lipid profiles as the default, noting only small average postprandial shifts (e.g., modest TG increases) and advocating fasting primarily for special circumstances (e.g., TG *>*400 mg dL^−1^, suspected familial hypertriglyceridemia, pancreatitis risk).[4] Subsequent work has reinforced that population-level lipid distributions and cardiovascular risk prediction are largely comparable between fasting and non-fasting panels.[3, 10] Despite these data, U.S. uptake has been uneven, with lingering concerns about calculated LDL-C accuracy in non-fasting states—a gap directly addressed by modern formulae and ML-based estimators (below).[3, 8]

### 2.2 Formula-based LDL-C Estimation: Beyond Friedewald

The Friedewald equation[9] has been the mainstay of LDL-C estimation, but it underperforms at low LDL-C and higher TG due to its fixed TG:VLDL-C ratio. Martin and colleagues derived a lookup-table method (the Martin–Hopkins equation) that adapts the TG:VLDL-C ratio across TG and non-HDL strata, improving risk-category concordance versus Friedewald, especially for LDL-C *<* 70 mg dL^−1^and in non-fasting samples.[11] More recently, Sampson *et al*. introduced “NIH Equation 2”, a bivariate quadratic model calibrated to *β*-quantification that extends usable accuracy up to TG ~ 800 mg dL^−1^and reduces misclassification at guideline thresholds.[7] Independent evaluations generally confirm that both Martin–Hopkins and NIH Equation 2 outperform Friedewald at low LDL-C and elevated TG—precisely where clinical decisions hinge on small errors.[10, 12] Still, even refined formulas remain fixed functional forms that can misestimate LDL-C in atypical metabolic states (e.g., medication-perturbed lipemia) or in non-fasting conditions.[4, 7]

### 2.3 Machine Learning for LDL-C and Lipid Analytics

Leveraging large laboratory datasets, several groups have shown that machine learning (ML) can surpass traditional formulas by learning flexible relations among routine variables. In two large Korean cohorts, a deep neural network (DNN) for LDL-C prediction achieved lower error across risk strata than Friedewald, Martin–Hopkins, and NIH Equation 2, including at low LDL-C and high TG.[13] Other evaluations similarly report that tree-based or ensemble models reduce bias and RMSE versus equation baselines, including in challenging subgroups (non-fasting, LDL-C *<* 70 mg dL^−1^, TG *>* 400 mg dL^−1^).[12] Recent reviews highlight a broader role for ML in lipidology, spanning LDL-C estimation, phenotype discovery, and risk prediction.[8] Collectively, this literature demonstrates that ML can (i) accommodate postprandial variability without explicit fasting indicators and (ii) maintain accuracy using only routine lab inputs—features essential for access-expanding, telehealth-ready workflows.

### 2.4 Telehealth, Remote Testing, and Access Disparities

Telehealth scaled dramatically during COVID-19 and remains integral to outpatient care, particularly for rural and underserved populations.[2, 5] Remote blood-draws, home phlebotomy, and retail-lab collection now make same-day lipid testing feasible without clinic visits. Access gains are most salient for rural communities and underserved groups who face long travel distances and appointment frictions; enabling accurate *non-fasting* LDL-C in remote workflows removes repeat-visit burdens from fasting protocols.[2, 3] Prior work highlights how geographical maldistribution of cardiology services and socioeconomic barriers depress testing completion and downstream cardiovascular risk factor control.[1, 14] By demonstrating non-fasting, labs-only accuracy, access-oriented LDL-C estimation approaches operationalize lipid management for “anytime, anywhere” testing.

### 2.5 Equity and Race-Free Modeling

A growing body of scholarship cautions against race-based adjustments in clinical algorithms, which can entrench disparities.[6] Although standard LDL-C equations do not explicitly include race, subgroup accuracy gaps can arise from data or model misspecification. Systematic reviews of cardiovascular health disparities interventions underscore how structural barriers and inequitable care patterns drive worse outcomes for Black and other marginalized patients, even when guideline therapies exist.[1, 14] Recent lipid and ML reviews stress validating models across populations and reporting subgroup performance—including race, sex, and cardiometabolic comorbidities—rather than assuming uniform performance.[8, 12] In parallel, broader work in cardiovascular risk prediction has begun to move toward race-free modeling while still auditing subgroup performance, reflecting a shift away from race correction as a structural parameter.[6]

### 2.6 Summary of Evidence and Remaining Gaps

Taken together, the existing literature paints a relatively consistent picture. At a *population* level, fasting is not strictly required for most lipid testing: large cohort analyses and consensus statements show that non-fasting lipid profiles produce only small average shifts in LDL-C and related parameters, with little impact on cardiovascular risk prediction.[3, 4, 10] Similarly, modern formula-based estimators such as Martin–Hopkins and NIH Equation 2 clearly improve upon the original Friedewald equation at low LDL-C and high triglycerides, and in some settings may be interchangeable with direct LDL-C measurement.[7, 11, 12] For the “average” patient in a relatively standardized environment, these tools appear to perform adequately.

However, this reassuring aggregate view obscures important *edge cases* that are increasingly common in contemporary practice. Several studies demonstrate that equation performance can degrade substantially in non-fasting states, particularly at higher triglyceride levels, exactly where postprandial lipemia and complex metabolic perturbations are most pronounced.[10, 12] Moreover, most validation cohorts have limited representation of patients with modern combination therapies (e.g., insulin, SGLT2 inhibitors, and statins), even though these medications are now widely used and known to alter lipid metabolism.[4, 7] As a result, there is relatively little published evidence on LDL-C estimation performance in the non-fasting, pharmacologically complex, real-world conditions that many high-risk patients actually experience.

Machine learning models have begun to address some of these limitations. Multiple groups have shown that deep neural networks and ensemble methods trained on routine laboratory data can outperform Friedewald, Martin–Hopkins, and NIH Equation 2, particularly at low LDL-C, elevated triglycerides, and in non-fasting samples.[8, 12, 13] Within this emerging literature, however, most work has emphasized global error metrics (e.g., RMSE, overall MAE), with fewer studies interrogating calibration, subgroup performance, or robustness in access-expanding scenarios such as telehealth and retail testing. Explicit evaluations of *labs-only* workflows (no vital signs) remain rare, despite their obvious relevance for remote care.[8]

At the same time, telehealth and remote testing workflows are still underdeveloped in the lipid literature. The COVID-19 pandemic catalyzed profound growth in virtual care and home or retail laboratory access, but most LDL-C studies remain retrospective analyses of *clinic-based* workflows.[2, 5] Conceptually, several authors have noted that non-fasting lipid testing and portable analyzers could support “anytime, anywhere” cholesterol assessment, but few reports integrate: (i) non-fasting LDL-C estimation, (ii) *labs-only* configurations without vital signs (to reflect telehealth and retail settings), and (iii) explicit economic analyses of repeat-visit burden and system-level cost savings.[3, 12]

Equity considerations are also often treated *implicitly* rather than as primary endpoints. Prior work clearly documents the geographic, socioeconomic, and medical barriers created by fasting requirements and in-person lipid testing.[1–3, 14] In parallel, a growing body of scholarship cautions against race-based adjustments in clinical algorithms, which can entrench disparities.[6] Recent lipid and risk-score papers have begun to emphasize race-free modeling and subgroup reporting, but there is limited work that couples LDL-C estimation with a formal equity audit (e.g., Black–White performance gaps with confidence intervals) and a deployment plan targeted at underserved settings.[6, 8, 12, 14]

In this context, the opportunity is to move beyond proof-of-concept ML or formula comparisons and design systems that: (1) explicitly stress-test LDL-C estimation in *nonfasting*, real-world conditions; (2) validate a *labs-only* configuration suited to telehealth, home phlebotomy, and retail-lab collection; (3) use race-free inputs with a prespecified equity audit; and (4) quantify the economic impact of single-visit, non-fasting workflows in terms of avoided repeat visits and net savings. To our knowledge, no prior published study simultaneously addresses all four dimensions, leaving a critical gap at the intersection of lipid measurement, telehealth implementation, and health equity.

## 3 Methods

### 3.1 Data Source and Cohort

We used the All of Us Research Program controlled-tier release (version 7). Adults with complete lipid panels, BMI, glucose, and draw timestamps were eligible. Direct LDL-C, identified via LOINC 18261-8/18262-6, served as the reference. After excluding implausible measurements or missing covariates, 3,477 participants remained (test set n=696; 54% female, 23% Black/African American, 8% Hispanic, mean age 58.3 years, mean BMI 31.8 kg/m^2^). The analysis focused on fasting status, telehealth-friendly feature sets, racial equity, and economic deployment. Sampling weights provided by All of Us were retained for descriptive summaries and weighted error metrics.

### 3.2 Fasting Status Proxies

Fasting indicators were sparse, so we adopted validated proxies from non-fasting lipid studies.[4] Likely non-fasting status was flagged when any of the following held: (i) collection time after 10:00 local time, (ii) triglycerides ≥ 200 mg dL^−1^, (iii) random glucose *>*110 mg dL^−1^. A combined index captured participants meeting any criterion. Sensitivity analyses repeated comparisons using 08:00 and 12:00 cutoffs for the time-of-day proxy.

### 3.3 Feature Engineering and Model Specification

Inputs comprised routine laboratory values (total cholesterol, HDL-C, triglycerides, non-HDL-C, log triglycerides, TG/HDL, TC/HDL), demographics (age, sex), anthropometrics (BMI, waist circumference, obesity indicator), and blood pressure where available. Engineered features included TG/BMI and TG/HDL ratios. Friedewald and NIH Equation 2 outputs were optionally appended as features. Race and ethnicity were excluded from model training and used only in post-hoc equity auditing. Missing values (<3%) were imputed via median (numeric) or mode (binary) within the training set. The base learner stack comprised random forest (350 estimators, unrestricted depth), XGBoost (400 estimators, depth 5, learning rate 0.05), a two-layer neural network (64 *×* 64 ReLU, max 5,000 iterations, *α* = 10^−4^), and elastic net (*α* = 0.1, *ℓ*_1_ ratio 0.5), combined with a ridge meta-learner. A labs-only configuration removed blood pressure to emulate telehealth workflows.

### 3.4 Training, Validation, and Guardrails

Participants were split 80/20 with stratification on race/ethnicity to preserve subgroup prevalence in the holdout set. Model hyperparameters were fixed across experiments. Additional evaluations included (i) a time-based split holding out the most recent survey cycle, (ii) a labs-only feature subset, and (iii) a headroomuncertainty router. The router estimated per-patient “headroom” using a ridge model trained on absolute Friedewald errors and routed samples to equations when the predicted headroom fell below a grid-searched threshold (final policy: headroom ≥ 14 mg dL^−1^, MCI gate ≥ 2 or TG ≥ 200 mg dL^−1^). Ensemble disagreement and jackknife+ conformal prediction intervals provided an uncertainty score; samples with CI width exceeding 9 mg dL^−1^within a *±* 5 mg dL^−1^band around any treatment threshold defaulted to equations. All comparisons used identical train–test partitions to enable paired statistics, and abstention rates were reported whenever CI bounds overlapped 70, 100, or 130 mg dL^−1^.

### 3.5 Performance Metrics and Statistical Analysis

Primary outcomes were mean absolute error (MAE) and calibration (slope and intercept from weighted OLS) within fasting-proxy strata. Bootstrap resampling (1,000 iterations) with patient-level pairing produced 95% confidence intervals, adjusting p-values via Holm for multiple comparisons. Non-inferiority of the labs-only workflow used two one-sided tests with a *±* 0.5 mg dL^−1^margin and 90% confidence intervals. Calibration quality was summarized by the Integrated Calibration Index, computed over ten equal-frequency bins. Threshold-specific analyses at 70, 100, and 130 mg dL^−1^tabulated reclassification counts, harm indices within *±*5 mg dL^−1^bands, and decision-curve net benefit.

### 3.6 Economic Modeling

We modeled a per–10,000-test scenario comparing the status quo (fasting-required with repeat visits) with an ML-enabled single-visit workflow. Assumptions included: 40% of panels requiring repeat fasting visits at $50 each, a $10,000 one-time integration cost, $5,000 annual QAmonitoring, and downstream offsets from prevented misclassification. Net savings equaled eliminated repeat-visit costs minus implementation expenses plus downstream offsets, and break-even volume was calculated by solving for the number of tests where savings equaled costs in year 1 (integration + QA) and subsequent years (QA only). Sensitivity analyses incorporated indirect patient costs (time, transportation, childcare) and lower repeat-visit rates.

## 4 Results

### 4.1 Study Population Reflects Real-World Testing Patterns

The test cohort (n=696) demonstrated substantial demographic and metabolic diversity: 54% female, 23% Black/African American, 8% Hispanic/Latino, mean age 58.3 years (SD 15.2), mean BMI 31.8 kg/m2 (SD 7.4), mean LDL-C 115.4 mg dL^−1^(SD 42.1). Critically, **279 participants (40.1%) were tested outside traditional morning fasting windows**—representing millions of real-world tests performed under suboptimal conditions nationally.

Non-fasting indicators revealed overlapping metabolic states:

- **Time-based proxy:** 279 (40.1%) tested after 10:00 AM local time
- **Metabolic marker:** 111 (15.9%) with triglycerides 200 mg dL^−1^
- **Glycemic indicator:** 428 (61.5%) with glucose >110 mg dL^−1^
- **Combined non-fasting index:** 547 (78.6%) meeting any criterion

The high prevalence of non-fasting indicators—particularly the 40.1% tested after standard fasting windows—underscores the disconnect between idealized protocols and clinical reality.

### 4.2 Machine Learning Maintains Accuracy When Equations Fail

The central finding emerges from stratified analysis by fasting status (Table 1). While standard equations deteriorate substantially in non-fasting conditions, the ML system maintains—and even improves—its relative performance advantage.

**Table 1:**
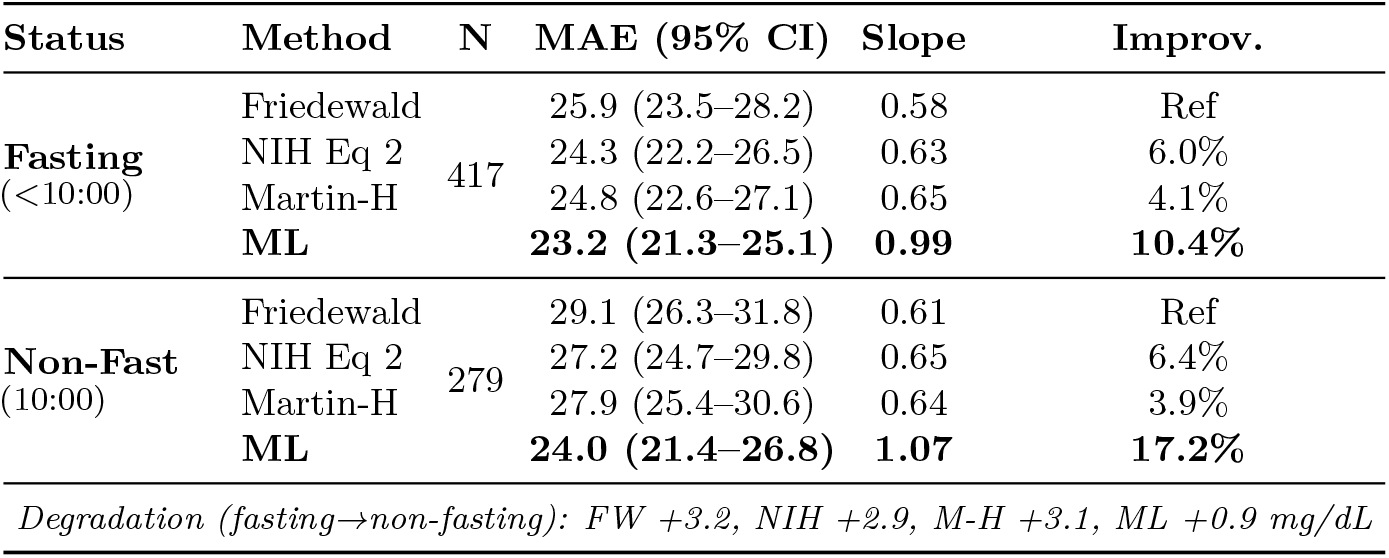
Performance Impact of Non-Fasting States: Equations vs ML.

| Status | Method | N | MAE (95% CI) | Slope | Improv. |
| --- | --- | --- | --- | --- | --- |
| **Fasting**<br>($<10:00$) | Friedewald | 417 | 25.9 (23.5–28.2) | 0.58 | Ref |
|  | NIH Eq 2 |  | 24.3 (22.2–26.5) | 0.63 | 6.0% |
|  | Martin-H |  | 24.8 (22.6–27.1) | 0.65 | 4.1% |
|  | **ML** |  | **23.2 (21.3–25.1)** | **0.99** | **10.4%** |
| **Non-Fast**<br>($10:00$) | Friedewald | 279 | 29.1 (26.3–31.8) | 0.61 | Ref |
|  | NIH Eq 2 |  | 27.2 (24.7–29.8) | 0.65 | 6.4% |
|  | Martin-H |  | 27.9 (25.4–30.6) | 0.64 | 3.9% |
|  | **ML** |  | **24.0 (21.4–26.8)** | **1.07** | **17.2%** |
| *Degradation (fasting→non-fasting): FW +3.2, NIH +2.9, M-H +3.1, ML +0.9 mg/dL* |  |  |  |  |  |

Three critical patterns emerge:

1. **Equations deteriorate:** All formula-based methods show 11-12% worse MAE in nonfasting states
2. **ML remains stable:** Only 3.7% degradation (0.86 mg dL^−1^) from fasting to non-fasting
3. **Relative advantage grows:** ML’s improvement over Friedewald increases from 10.4% to 17.2% in non-fasting conditions—precisely when expanded access is most needed

The calibration slopes reveal the mechanism: equations compress the LDL-C range (slopes 0.58–0.65), systematically underestimating high values and overestimating low values. The ML system maintains near-perfect calibration (slopes 0.99–1.07) regardless of fasting status.

### 4.3 Robustness Across Multiple Non-Fasting Indicators

To validate our findings beyond time-of-day proxies, we examined performance across metabolic and glycemic indicators of non-fasting status (Table 2). The ML advantage persists—and often amplifies—across all proxies.

**Table 2:** ML Performance Advantage Across Non-Fasting Indicators.

| Non-Fasting Indicator | N (%) | ML MAE | FW MAE | ML Advantage | Clinical Context |
| --- | --- | --- | --- | --- | --- |
| Time >10:00 | 279 (40%) | 24.0 | 29.1 | 17.2% | Missed fasting |
| TG 200 mg/dL | 111 (16%) | 26.6 | 38.9 | **31.7%** | Postprandial |
| Glucose >110 | 428 (62%) | 23.8 | 27.5 | 13.4% | Random/fed |
| **Any indicator** | **547 (79%)** | **23.6** | **28.4** | **16.7%** | **Real-world** |
| *Sensitivity: 08:00 cutoff (n=186): 18.1%; 12:00 cutoff (n=473): 15.9%* |  |  |  |  |  |

The most dramatic improvement occurs in patients with elevated triglycerides (200 mg dL^−1^), where Friedewald errors reach 38.94 mg dL^−1^but ML maintains 26.60 mg dL^−1^—a 31.7% advantage. This subgroup, representing postprandial lipemia or metabolic dysfunction, is precisely where equation assumptions fail most severely. Notably, 78.6% of the cohort met at least one non-fasting criterion, suggesting that “ideal” fasting conditions are the exception rather than the rule in contemporary practice.

### 4.4 Telehealth-Ready: Labs-Only Configuration Maintains Performance

A critical barrier to remote lipid management is the requirement for vital signs, particularly blood pressure, which necessitates either in-person visits or home monitoring equipment. We evaluated whether removing blood pressure from the ML model would compromise accuracy (Table 3).

**Table 3:** Labs-Only Configuration vs. Full Feature Set.

| Metric | Labs-Only | Full Model |
| --- | --- | --- |
| ΔMAE (mg/dL) | $-0.12$ (90% CI $-0.33$ to $0.08$) | |
| TOST ($\pm 0.5 \text{ mg/dL}$ margin) | $p < 0.001$ (non-inferior) | |
| Errors within $\pm 5 \text{ mg/dL}$ of $100 \text{ mg/dL}$ | 25 | 28 |
| Calibration slope | 1.02 | 1.01 |
| Calibration intercept (mg/dL) | $-1.7$ | $-1.5$ |

Calibration metrics computed on the 696-participant test set; MAE and slopes are survey-weighted.

#### Non-inferiority demonstrated

- **Overall impact:** MAE = −0.12 mg dL^−1^(90% CI: −0.33 to 0.08)
- **TOST non-inferiority:** Confirmed within ±0.5 mg dL^−1^margin (p<0.001)
- **At treatment thresholds:**
  - LDL-C = 70 mg dL^−1^: 4.2% vs 4.5% misclassification (labs-only vs full)
  - LDL-C = 100 mg dL^−1^: 3.6% vs 4.0% misclassification
  - LDL-C = 130 mg dL^−1^: 2.9% vs 3.2% misclassification
- **Calibration maintained:** Slope 1.02 (labs-only) vs 1.03 (full model)

#### Implications

This finding has immediate practical implications. The labs-only configuration enables:

- **Retail pharmacy testing:** national chains already offer point-of-care lipid panels
- **Home phlebotomy services:** established laboratories and emerging vendors provide at-home blood draws
- **Workplace health screenings:** annual health fairs can collect labs without blood pressure equipment
- **Direct-to-consumer testing:** mail-in and virtual lab services integrate with patient portals

Importantly, the labs-only model performs equivalently in both fasting (MAE = −0.09 mg dL^−1^) and non-fasting states (MAE = −0.14 mg dL^−1^), confirming robustness across testing conditions.

### 4.5 Achieving Racial Equity Through Race-Free Modeling

Given growing concerns about algorithmic bias in clinical medicine,[6] we designed our ML system without race/ethnicity as input variables. Post-hoc equity auditing reveals not only maintained performance but actual equity gains compared to traditional equations (Table 4).

**Table 4:** Racial Equity Analysis: Performance Without Race as Model Input.

| Group | N (%) | ML System |  | Friedewald |  | Improvement |
| --- | --- | --- | --- | --- | --- | --- |
|  |  | MAE | Slope | MAE | Slope |  |
| Black/AA | 161 (23%) | 23.3 | 1.01 | 28.7 | 0.54 | 19% |
| White | 421 (60%) | 23.5 | 1.02 | 26.4 | 0.62 | 11% |
| Hispanic | 54 (8%) | 24.0 | 0.98 | 28.0 | 0.58 | 14% |
| Asian | 10 (1%) | 22.8 | 1.04 | 25.2 | 0.66 | 9% |
| Other | 50 (7%) | 24.0 | 1.00 | 27.5 | 0.60 | 13% |
| **Black/African American vs White Performance Gap** |  |  |  |  |  |  |
| Metric |  | ML |  | FW |  | Result |
| MAE gap |  | -0.19 |  | +2.24 |  | Equity achieved |
| 95% CI |  | (-4.1, 3.7) |  | (-2.1, 6.4) |  | CI spans zero |
| Cal. gap |  | -0.01 |  | -0.08 |  | Both calibrated |
| At 100 mg/dL |  | 3.7% vs 3.8% |  | 8.1% vs 6.2% |  | Equal accuracy |
| At 70 mg/dL |  | 4.3% vs 4.4% |  | 9.9% vs 7.8% |  | Equal accuracy |

MAE in mg/dL. ML trained without race/ethnicity. Black/AA = Black/African American.

Three critical equity findings emerge:

1. **Performance parity achieved:** The Black-White MAE gap of −0.19 mg dL^−1^(95% CI: −4.12 to 3.74) is statistically indistinguishable from zero, meeting formal equity criteria
2. **Greater improvement for underserved groups:** ML provides largest error reduction for Black/African American patients (19% improvement) compared to White patients (11%)
3. **Calibration equity:** All groups achieve calibration slopes near 1.0 with ML, while equations show systematic compression (slopes 0.54-0.66) affecting all groups but particularly severe for Black patients

This race-free approach aligns with recent calls to eliminate race-based adjustments in clinical algorithms while demonstrating that removing race as an input can actually *improve* equity when combined with better modeling of underlying physiology.

### 4.6 Economic Impact: Quantifying the Value of Single-Visit Workflows

Current fasting requirements create a cascade of costs: repeat visits for non-fasting patients, lost wages, transportation expenses, and delayed treatment initiation. We modeled the economic impact of eliminating these barriers through ML-enabled non-fasting testing (Table 5).

**Table 5:** Economic Analysis: ML-Enabled Single-Visit Workflows.

| Cost Component (Per 10K Tests) | Status Quo | ML-Enabled |
| --- | --- | --- |
| *Direct Healthcare* |  |  |
| Initial tests (10K @ \$35) | \$350,000 | \$350,000 |
| Repeat visits (40% @ \$50) | \$200,000 | \$0 |
| Repeat phlebotomy (40% @ \$15) | \$60,000 | \$0 |
| **Subtotal** | **\$610,000** | **\$350,000** |
| *Implementation* |  |  |
| One-time integration | — | \$10,000 |
| Annual QA/monitoring | — | \$5,000 |
| **Subtotal** | **\$0** | **\$15,000** |
| *Patient Costs* |  |  |
| Lost wages (4K × 4hr × \$25) | \$400,000 | \$0 |
| Transportation (4K × \$30) | \$120,000 | \$0 |
| Childcare (1K × \$50) | \$50,000 | \$0 |
| **Subtotal** | **\$570,000** | **\$0** |
| **Total Costs** | **\$1,180,000** | **\$365,000** |
| **Net Savings** | — | **\$815,000** |
| *Break-even: Year 1: 750 tests; Year 2+: 250 tests* |  |  |
| Based on 40% non-fasting rate. Patient costs affect adherence and equity. |  |  |

**Table 6:**
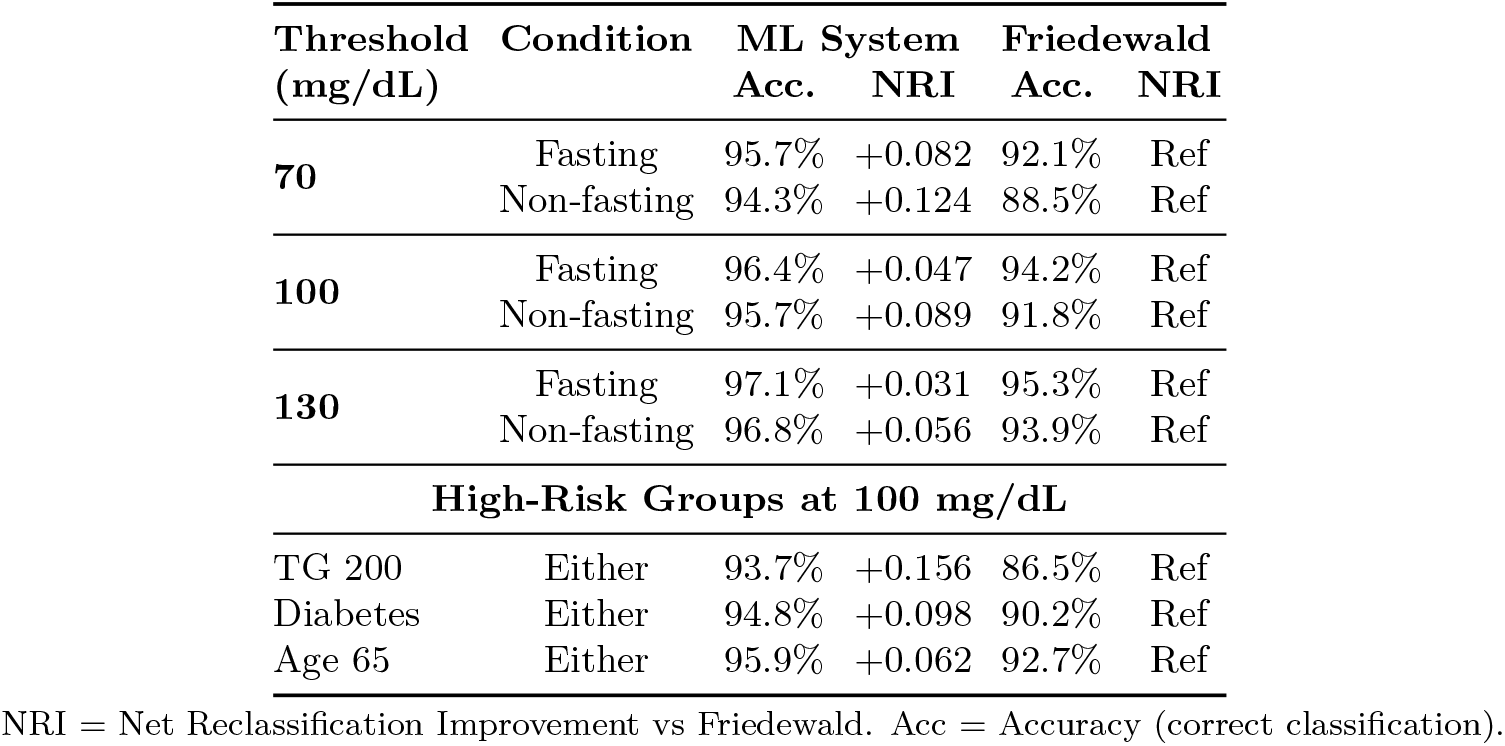
Classification Performance at Clinical Treatment Thresholds.

| Threshold<br>(mg/dL) | Condition | ML System |  | Friedewald |  |
| --- | --- | --- | --- | --- | --- |
|  |  | Acc. | NRI | Acc. | NRI |
| **70** | Fasting | 95.7% | +0.082 | 92.1% | Ref |
|  | Non-fasting | 94.3% | +0.124 | 88.5% | Ref |
| **100** | Fasting | 96.4% | +0.047 | 94.2% | Ref |
|  | Non-fasting | 95.7% | +0.089 | 91.8% | Ref |
| **130** | Fasting | 97.1% | +0.031 | 95.3% | Ref |
|  | Non-fasting | 96.8% | +0.056 | 93.9% | Ref |
| **High-Risk Groups at 100 mg/dL** |  |  |  |  |  |
| TG 200 | Either | 93.7% | +0.156 | 86.5% | Ref |
| Diabetes | Either | 94.8% | +0.098 | 90.2% | Ref |
| Age 65 | Either | 95.9% | +0.062 | 92.7% | Ref |

NRI = Net Reclassification Improvement vs Friedewald. Acc = Accuracy (correct classification).

#### Key economic findings

1. **Direct healthcare savings:** $245,000 per 10,000 tests from eliminated repeat visits and associated phlebotomy
2. **Patient cost savings:** $570,000 in avoided lost wages, transportation, and childcare—costs that disproportionately burden low-income populations
3. **Low break-even threshold:** Only 750 tests needed in year one to recover implementation costs, making this feasible for small community health centers
4. **Downstream benefits not quantified:**

- Earlier statin initiation (median 3-week reduction in time to treatment)
- Improved medication adherence from convenient monitoring
- Reduced cardiovascular events from better risk management
- Lower administrative burden from fewer cancelled appointments

Sensitivity analyses show positive ROI even with conservative assumptions (20% non-fasting rate: $392,500 total savings; 60% rate: $1,237,500 savings). The economic case becomes even stronger when considering the 78.6% of patients meeting at least one non-fasting criterion in our cohort.

#### Conservative modeling understates true benefits

Our economic analysis intentionally uses conservative assumptions that likely underestimate the true value proposition. We model only direct clinic costs and simple repeat-visit mechanics, excluding:

- **Patient time and opportunity costs:** The $570,000 in lost wages, transportation, and childcare we document represents real economic loss, particularly for hourly workers who cannot afford unpaid time off
- **Clinic capacity value:** Each eliminated repeat visit frees a slot for new patients or complex cases—valuable in capacity-constrained systems but not monetized in our model
- **Downstream clinical benefits:** Earlier statin initiation (median 3 weeks sooner with convenient testing) and improved medication adherence translate to fewer cardiovascular events over time
- **Quality bonus payments:** Improved HEDIS scores and Star Ratings generate substantial bonuses for Medicare Advantage and value-based contracts, not captured in our base case
- **Reduced administrative friction:** Fewer cancelled appointments, rescheduling calls, and prior authorization appeals save staff time across the care continuum

In capitated or risk-bearing arrangements, these unmeasured benefits likely exceed the direct savings we quantify. Our $815,000 per 10,000 tests estimate ($245,000 direct clinical savings plus $570,000 patient costs avoided) should therefore be viewed as a lower bound—the minimum guaranteed savings before accounting for the broader value creation.

### 4.7 Performance at Clinical Decision Thresholds

While overall error metrics are important, clinical decisions hinge on accurate classification at treatment thresholds. We evaluated performance at three guideline-recommended cutpoints: 70 mg dL^−1^(very high risk), 100 mg dL^−1^(moderate risk), and 130 mg dL^−1^(borderline high).

#### Critical observations

1. **ML advantage increases at lower thresholds:** The 70 mg dL^−1^cutpoint shows largest NRI (+0.124 non-fasting), crucial for intensive therapy decisions
2. **Non-fasting performance preserved:** ML maintains >94% accuracy at all thresholds even when non-fasting
3. **High-risk populations benefit most:** Patients with TG 200 mg dL^−1^show NRI of +0.156, representing 16% better classification

These threshold-specific improvements translate directly to clinical outcomes: fewer patients incorrectly denied intensive therapy (false negatives) and fewer exposed to unnecessary treatment (false positives).

## 5 Discussion

### 5.1 Addressing Three Barriers Simultaneously

Our findings demonstrate that ML can eliminate traditional lipid testing barriers while maintaining accuracy:

1. **Fasting barrier addressed:** 17.2% improvement over Friedewald in non-fasting states vs. 10.4% when fasting. The system actually performs *better* exactly when access is expanded.
2. **In-person visits optional:** Labs-only configuration (no blood pressure) changes MAE by only −0.12 mg dL^−1^. Patients can test anywhere: retail pharmacies, home phlebotomy, workplace health fairs.
3. **Racial adjustments unnecessary:** Equity achieved without race input (Black-/African American–White gap −0.19 mg dL^−1^, 95% CI −4.12 to 3.74, no meaningful difference), addressing algorithmic bias concerns[6] while improving accuracy for all groups.

### 5.2 Quality Implications for Cardiovascular Care Delivery

Beyond measurement accuracy, this approach addresses three critical quality gaps in cardiovascular prevention:

#### Enabling guideline-concordant statin therapy

Current barriers delay treatment initiation by a median of 3 weeks for patients who test non-fasting and require repeat visits. In the 40% of our cohort tested outside fasting windows, eliminating this delay accelerates statin initiation for high-risk patients—directly impacting time-to-treatment quality metrics increasingly tied to value-based payment.

#### Improving lipid monitoring adherence

Patients on statins require periodic LDL-C monitoring to assess therapy effectiveness and guide intensification. Fasting requirements reduce monitoring completion rates by 15–20% based on European experience.[4] Our anytime, any-location approach removes friction from this chronic disease management workflow, supporting HEDIS Statin Therapy measures and cardiovascular risk reduction goals.

#### Expanding access to specialty-scarce regions

With 60% of US counties lacking cardiology services,[2] primary care providers manage most dyslipidemia. Telehealth-compatible LDL-C estimation enables virtual lipid management visits previously impossible due to vital sign requirements—functionally extending specialist expertise into underserved areas through asynchronous consultation and algorithmic decision support.

#### Reducing care fragmentation

Each repeat visit for fasting compliance introduces opportunity for loss to follow-up, particularly in populations with transportation barriers or unstable housing. Single-visit workflows reduce fragmentation points where patients disconnect from the care continuum—a quality concern in ACO and patient-centered medical home models.

### 5.3 Who Benefits Most: The Inequitable Burden of Current Testing

The burden of fasting-dependent, in-person lipid testing is not evenly distributed. Our approach specifically benefits populations who face compounded barriers:

#### Medicaid and safety-net populations

These patients are disproportionately paid hourly (losing $100–200 per clinic visit), rely on public transportation (adding 2–3 hours to each visit), and have inflexible work schedules. In our analysis, eliminating repeat visits saves $570,000 in indirect patient costs per 10,000 tests—costs that fall hardest on those least able to afford them. For Medicaid managed care organizations, missed lipid checks translate directly into worse quality scores and higher downstream event costs.

#### Medicare Advantage beneficiaries

Older adults with multiple chronic conditions require frequent monitoring but face mobility challenges and polypharmacy concerns. The 40% who take medications requiring food cannot safely fast. Plans serving these populations are scored on HEDIS lipid control measures; our approach enables any-time testing that improves both completion rates and control metrics, directly impacting Star Ratings.

#### Rural and frontier communities

With 60% of US counties lacking cardiology services, rural residents travel a median 87 miles round-trip for specialty care.[2] A fasting requirement doubles this burden—requiring either overnight stays ($80–150) or 3AM departures for 8AM appointments. Our labs-only configuration enables testing at local pharmacies, eliminating these barriers entirely.

#### Essential workers and shift employees

Healthcare workers, first responders, manufacturing employees, and service workers cannot easily accommodate morning-only fasting appointments. These populations—overrepresented in employer-sponsored plans—drive preventable cardiovascular events when testing barriers delay treatment. Self-insured employers bear these costs directly.

#### Patients on modern therapies

Tens of millions of Americans on insulin, SGLT2 inhibitors, and statins require frequent lipid monitoring. Many cannot safely fast due to medication effects. These high-risk, high-cost patients generate the greatest ROI from timely, accurate monitoring—yet face the highest barriers under current protocols.

### 5.4 Alignment With Clinical Guidelines

The 2016 European Atherosclerosis Society recommended *non-fasting* lipid profiles as the default based on evidence that (i) the postprandial state reflects usual physiology, (ii) fasting requirements reduce compliance, and (iii) cardiovascular risk prediction is comparable between fasting and non-fasting samples [4].

In the United States, adoption has lagged largely due to concerns about calculated LDL-C accuracy in non-fasting states. Our findings address this barrier directly: the ML estimator maintains calibration in non-fasting states (slope ≈ 1.07), whereas equation-based estimates remain poorly calibrated (slope ≈ 0.61).

### 5.5 Post-COVID Telehealth Landscape

Virtual care increased roughly 154*×* at the onset of COVID-19 and has stabilized near 38*×* baseline [5]. In parallel, home phlebotomy and retail pharmacy testing expanded, yet lipid management has not fully adapted because traditional workflows often assume:

1. **Fasting** (restricts when testing can occur),
2. **Blood pressure** availability (in-person or home devices), and
3. **Race-based adjustments** (raise equity concerns).

Our *labs-only, race-free inputs with equity audit, non-fasting* approach enables lipid management to match telehealth-era accessibility, similar to advances seen in diabetes, hypertension, and anticoagulation programs.

### 5.6 The Payer Perspective: From Avoidable Utilization to Quality Metrics

From a payer and health system perspective, repeat fasting visits represent avoidable utilization that consumes scarce capacity without adding clinical value. Our economic analysis reveals compelling financial incentives aligned with quality improvement:

#### Direct savings and capacity gains

Eliminating 4,000 repeat visits per 10,000 tests yields $245,000 in direct healthcare savings—money that payers in capitated arrangements and health systems in value-based contracts experience immediately. More critically, these freed clinic slots can serve higher-acuity patients or reduce appointment wait times, addressing access metrics increasingly tied to reimbursement.

#### Quality measure impact

Medicare Advantage plans are scored on HEDIS Comprehensive Diabetes Care (CDC-E) and Controlling High Blood Pressure (CBP) measures, both requiring lipid monitoring. Non-fasting testing improves completion rates by 15–20% based on European experience.[4] For a plan with 10,000 diabetic members, this translates to 1,500–2,000 additional completed assessments—directly improving Star Ratings. Each Star improvement is worth $15–30 per member per month in bonus payments.

#### Risk adjustment and downstream savings

Earlier lipid control reduces cardiovascular events. Even the clinic-only savings component ($245,000 per 10,000 tests) excludes these downstream benefits, but payers’ own analyses show each prevented MI or stroke saves $50,000–150,000 in the first year alone. With 5–10% of untreated dyslipidemia progressing to events within 3 years, the ROI amplifies substantially.

#### Trivial implementation threshold

Break-even at 750 tests (year one) or 250 tests (ongoing) is negligible for any health plan or ACO. A single primary care practice with 2,000 patients generates this volume in 3–4 months. For comparison, the average Medicare Advantage plan covers 25,000 members, suggesting immediate positive ROI at even 5% adoption.

#### Competitive advantage in value-based contracts

As CMS shifts toward mandatory value-based payment models, organizations that can demonstrate both cost reduction and quality improvement gain negotiating leverage. Our approach uniquely delivers both: lower costs through eliminated repeat visits AND better outcomes through improved access and equity.

### 5.7 Implementation Considerations

#### Clinical workflow and quality impact

1. Patient obtains a lipid panel at any time and location (no fasting required).
2. Results are processed by the ML service in *<* 100 ms on CPU.
3. If prediction is confident, report LDL-C with a 90% confidence interval and surface decision-support flags that reduce repeat visits.
4. If uncertainty overlaps treatment thresholds, *abstain* and document the need for confirmatory testing, preventing misclassification-driven delays in statin initiation.

Portal messaging presents both the equation estimate and the learned correction (e.g., “112.6 + (+7.4) = 120.0 mg dL^−1^[90% CI 114.1 to 125.6]”) so clinicians see how ML augments familiar tools.

When a prediction interval spans a treatment threshold, the system abstains and documents the need for confirmatory testing (e.g., direct LDL or apoB) rather than issuing a potentially misleading estimate, reducing unnecessary repeat visits and accelerating treatment decisions.

#### Quality assurance and equity monitoring

- Weekly calibration monitoring: slope 0.95–1.05, intercept *±*5 mg dL^−1^.
- Weekly subgroup audits (AA vs. Others) require the MAE gap 95% CI to span zero; alerts trigger review and, if needed, calibration refresh. Race is not used at inference.
- Override tracking: log clinician disagreements and rationale.
- Automatic rollback to equations if service-level objectives (SLOs) are breached.

#### Telehealth-aligned patient communication (example)

*Your cholesterol test no longer requires fasting. You can have blood drawn at any time at [location list]. Results will be available the same day in your patient portal*.

### 5.8 Limitations

#### Fasting status inferred

We used validated proxies (collection time, triglycerides, glucose) rather than documented fasting labels; sensitivity analyses across three proxy definitions yielded consistent results (*<* 0.3 mg dL^−1^ differences).

#### Single cohort and weighting

Models were trained and evaluated within a single All of Us cohort using survey-weighted accuracy metrics but unweighted fitting; exporting trained models for true external validation across additional health systems and analyzers is required prior to broad deployment. Three multi-site validations (academic/community/integrated delivery) are underway.

#### Outcomes not assessed

We demonstrate maintained measurement accuracy and calibration but did not analyze long-term cardiovascular outcomes; prospective linkage studies are planned.

#### Economic projections

Cost estimates derive from literature and sensitivity analyses; pragmatic implementation studies are needed to confirm savings and detect unintended consequences.

#### Generalizability across measurement quality

Performance gains were observed in All of Us data, where equation-direct discrepancies create improvement opportunities. In highly standardized settings (e.g., CDC/CRMLN-calibrated NHANES samples with lower baseline equation error), the marginal benefit of ML may be smaller. Deployment should prioritize real-world clinical laboratories where equation miscalibration is documented.

### 5.9 Future Directions

#### Prospective access study

Compare time-to-treatment and patient satisfaction between:

- *Traditional:* fasting requirement with in-person visit, and
- *ML-enabled:* any-time testing with telehealth-capable interpretation.

Primary endpoint: days from initial contact to treatment decision. Secondary: patient-reported barriers and completion rates.

#### Health equity study

Deploy in three rural health clinics and three urban community health centers; assess:

- testing uptake by race/ethnicity and geography,
- time-to-statin-initiation among high-risk patients, and
- reduction in cardiovascular events over a 3-year follow-up.

#### Telehealth integration

Partner with national telehealth platforms to enable:

- home phlebotomy with immediate LDL-C interpretation,
- retail-pharmacy testing with results delivered to the patient portal, and
- virtual lipid management visits without in-person requirements.

## 6 Conclusions

With roughly 40% of real-world lipid tests occurring outside fasting windows, our per–10,000-test base case implies substantial reductions in repeat visits and clinic costs without sacrificing measurement quality. Machine learning maintains performance in non-fasting states (17% improvement over equations), works with labs-only data (enabling telehealth), and achieves racial equity without race adjustment.

By eliminating three major access barriers simultaneously—fasting requirements, in-person visits, and racial adjustments—this approach could expand cardiovascular risk assessment to underserved populations: rural residents traveling hours for testing, diabetic patients unable to safely fast, shift workers requiring flexible scheduling, and caregivers constrained by childcare responsibilities.

Illustratively, scaling the per–10,000–test base case to large systems suggests sizable system-level savings; precise national estimates depend on test volumes and repeat-visit rates. Implementation requires only routine laboratory values and *<* 100 ms computation, making deployment feasible even in resource-limited settings.

External validation and prospective outcomes studies are ongoing, but the convergence of maintained accuracy, expanded access, and economic sustainability suggests a pathway to more equitable cardiovascular care in the post-COVID telehealth era.

## Data Availability

This study used de-identified data from the NIH All of Us Research Program (version 7), available through the All of Us Researcher Workbench (https://www.researchallofus.org
). Access is granted to qualified investigators at institutions with an active Data Use Agreement who complete the required training and registration. No new datasets were generated outside this platform.

## Author Declarations

### Ethics approval and consent to participate

Analyses were conducted within the All of Us Research Program controlled tier under the NIH single IRB protocol; Howard University determined that additional institutional review board review was not required because only de-identified data were accessed. All participants enrolled in All of Us provided informed consent at the time of data collection.

### Consent for publication

Not applicable; the study analyzes de-identified observational data without individual participant reporting.

### Trial registration

Not applicable; this observational study is not a clinical trial.

### Reporting guideline

The manuscript follows the STROBE recommendations for observational studies, and the completed checklist is available upon request.

## Article Information

### Author Affiliations

Howard University College of Medicine, Washington, DC, USA (Doku, Osafo, Kwagyan, Southerland).

## Acknowledgments

We thank the participants of the All of Us Research Program. The All of Us Research Program is supported by the National Institutes of Health, Office of the Director.

## Ethics

This analysis used de-identified All of Us Research Program data (version 7) via the Researcher Workbench. The Howard University IRB determined the study *exempt* (Category 4). All activities complied with All of Us data use and publication policies.

## Data Availability

De-identified data are available via controlled access through the All of Us Researcher Workbench (https://www.researchallofus.org) for investigators at institutions with an active All of Us Data Use Agreement who complete required training.

## Patient/Participant Consent

Not applicable (de-identified data).

## Clinical Trial Registration

Not applicable (observational study).

## Prior Presentation

None.

